# Research at the Intersection of Traditional, Complementary, and Integrative Medicine and Artificial Intelligence: A Bibliometric Analysis

**DOI:** 10.64898/2025.12.09.25341890

**Authors:** Henry Liu, Mirela-Ioana Bilc, Jeremy Y. Ng

## Abstract

**Background:** Traditional, complementary, and integrative medicine (TCIM) describes a broad collection of medical interventions, practices, and belief systems that fall outside the purview of conventional medicine. Accompanying the recent growth of TCIM research productivity, artificial intelligence (AI) technologies have increasingly impacted areas of biomedical research, including diagnosis, treatment planning, and drug discovery. This bibliometric analysis explores the characteristics of research publications at the intersection of TCIM and AI.

**Methods:** A search string encompassing terms related to TCIM and AI was run on MEDLINE on 14 November 2025, with no restrictions by date, language, or publication type. Retrieved MEDLINE records were subsequently used for DOI citation searches in Scopus, with results exported on the same date. The following bibliometric data were collected: number of publications (including total publications and publications per year), open access status, subject area, document type, publication stage, publications per journal, author affiliations; funding sponsors, publication country, source type, and publication language. Trends in the bibliographic were generated in Excel, and bibliometric networks were visualized using VOSviewer, with thematic clusters identified and presented.

**Results:** A total of 1917 publications (n = 921 open-access) by 9749 unique authors, were published from 1991 to 2026. The greatest number of publications were published over the last 5 years, with the most productive journals/sources being Scientific Reports (n = 86), PLoS One (n = 63), and IEEE Transactions on Neural Systems and Rehabilitation Engineering (n = 59). The most productive countries included China (n = 982), the United States (n = 353), and the United Kingdom (n = 90), with frequent institutional affiliations and funding sponsors also being from these countries.

**Conclusions:** This bibliometric analysis provides insights into research productivity at the intersection of the TCIM and AI, showing rapidly increasing growth in the field. Future work can continue to investigate changes in the publication characteristics of emerging research, as the volume of publications on this topic is expected to rapidly grow.

## Background

Traditional, complementary, and integrative medicine (TCIM) encompasses a broad spectrum of medical and healthcare practices that exist outside the framework of conventional Western medicine.^1^ These practices include, but are not limited to, herbal medicine, acupuncture, yoga, meditation, Ayurveda, traditional Chinese medicine (TCM), homeopathy, naturopathy, and various forms of mind-body interventions.^2,3^ The World Health Organization (WHO) recognizes TCIM as an essential component of healthcare in many countries, with over 170 member states reporting formal policies, laws, and regulations governing its practice.^4,5^ The global prevalence of TCIM use among patients is significant, with increasing acceptance of TCIM among conventional healthcare providers and growing integration into mainstream healthcare systems.^6–8^

Simultaneously, artificial intelligence (AI) is transforming various domains of healthcare, including diagnostics, treatment planning, drug discovery, and personalized medicine.^9–11^ AI encompasses a range of technologies such as machine learning (ML), deep learning (DL), natural language processing (NLP), and neural networks, which enable the automation of complex tasks, data analysis, and decision-making processes.^11,12^ In conventional medicine, AI has been leveraged to improve diagnostic accuracy, enhance medical imaging, optimize patient management, and facilitate biomedical research.^10,13–15^ As AI technologies continue to evolve, their application in TCIM research and practice presents a promising frontier for innovation.^4,16^

Despite the rapid advancement of AI in healthcare, the intersection of AI and TCIM remains relatively underexplored.^4,16,17^ Existing literature suggests that AI has potential applications in TCIM, including predictive modeling of treatment outcomes, optimization of herbal medicine formulations, digital monitoring of TCIM therapies, and the development of AI-assisted diagnostic tools for traditional medicine-based approaches.^16,18–21^ For example, machine learning models have been used to analyze vast datasets of herbal compounds to identify potential therapeutic properties, while NLP techniques have been employed to extract insights from classical TCIM texts.^22,23^ Additionally, technologies powered by AI are increasingly being utilized to track the physiological effects of interventions such as acupuncture, yoga, and meditation, providing objective data to support their efficacy and safety.^20,24,25^ AI has the potential to bridge gaps in TCIM research by providing evidence-based insights, improving standardization, and enhancing clinical decision-making.^26,27^

However, one of the primary challenges in studying the intersection of AI and TCIM is a lack of standardized methodologies and integration frameworks. In contrast to conventional medicine, TCIM encompasses diverse and often heterogeneous practices rooted in different cultural, historical, and philosophical traditions.^1,28^ The application of AI in TCIM therefore requires interdisciplinary collaboration between computer scientists, healthcare practitioners, and researchers to ensure that technological advancements align with the principles and holistic nature of TCIM.^16^ Furthermore, ethical considerations, such as data privacy, informed consent, and the potential biases embedded in AI algorithms, must be carefully addressed to ensure responsible and equitable implementation.^29–32^ Given the increasing interest in both AI and TCIM, understanding the existing body of literature at this intersection could help future researchers, policymakers, and practitioners navigate the opportunities and challenges associated with integrating AI into TCIM.^4,16,33,34^

Bibliometric analysis is a quantitative approach to assessing scientific literature, providing insights into publication trends, citation networks, authorship patterns, and thematic foci.^35–37^ Previous bibliometric studies have analyzed TCIM research and AI applications separately^33,34^; however, to date, no comprehensive analysis has been conducted to explore the intersection of the whole fields of AI and TCIM. A bibliometric analysis of the research landscape at the intersection of TCIM and AI is essential to map the current state of knowledge, identify key contributors, and highlight emerging trends and research gaps.

## Methods

### Study Design

This study is a bibliometric analysis that systematically examines the characteristics of research publications at the intersection of TCIM and AI. The study was designed in accordance with best practices in bibliometric research and follows existing preliminary guidance for transparent reporting of bibliometric analyses.^38, 39^ The study was registered on the Open Science Framework (OSF) at https://doi.org/10.17605/OSF.IO/3UJXZ to ensure methodological transparency and reproducibility.

### Data Sources and Software

The primary data source for this bibliometric analysis was MEDLINE via the Ovid platform, one of the largest and most comprehensive abstract and citation databases of peer-reviewed literature. All retrieved articles were exported with their digital object identifiers (DOIs). Articles’ DOIs were then used to perform citation searches in Scopus to identify publications that cited the original MEDLINE records, enabling the construction of a comprehensive citation dataset for further analysis. MEDLINE and Scopus searches were both performed on November 14, 2025 to prevent discrepancies between database updates.

The following bibliometric data were collected from Scopus: number of publications (including total publications and publications per year), open access status, subject area, document type, publication stage, publications per journal, author affiliations; funding sponsors, publication country, source type, and publication language. Impact factors were manually searched based on Journal Citation Reports (JCR) for the ten journals with the highest number of publications.

All statistical analyses were conducted using Excel and VOSviewer. Descriptive statistics were generated using Excel. Network analyses and visualizations were generated using VOSviewer based on Scopus search results, allowing the identification of co-authorship networks, keyword co-occurrence, and thematic trends within the field.

### Search Strategy

A structured MEDLINE search strategy was developed to identify publications, including keywords and Medical Subject Headings (MeSH) terms related to TCIM and AI, combined using Boolean operators (**Table 1**). The search strategy was developed by a methodologist (JYN) with over a decade of experience developing search strategies in the TCIM field. The search was executed to capture records of all relevant publications from inception to the date of last database update. To ensure search reproducibility, complete search strategies, including database queries and number of results, are provided in **Appendix 1** on OSF: https://osf.io/cpmgu/files/zc36d.

**Table 1:**
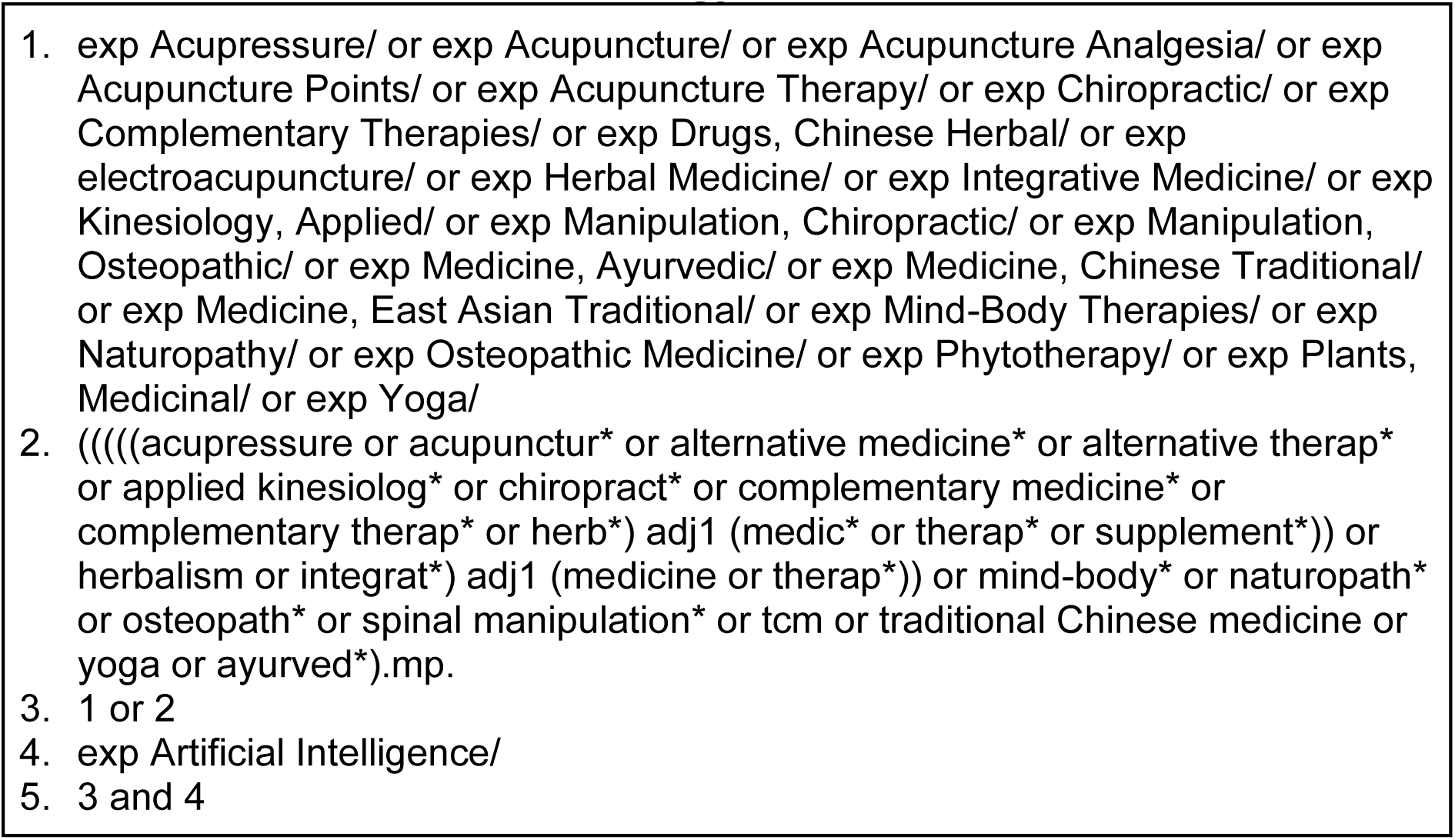
MEDLINE Search Strategy.

### Bibliometric Analysis

Descriptive statistics were generated to summarize publication trends and the general characteristics of publications, including annual growth in the number of publications, document and source types, subject areas of publication, languages of publication, and author characteristics such as associated country and institutional affiliations. The most highly cited articles and most productive sources were identified and characterized. Co-authorship analysis was performed to examine collaboration patterns between countries, where VOSviewer was used to generate visual representations of this network. A co-occurrence analysis of author keywords was conducted using VOSviewer to identify key research themes and emerging trends, with keywords were grouped into clusters to visualize topic distributions at the intersection of TCIM and AI research. A co-citation analysis of the most prolific sources was also conducted using VOSviewer, identifying co-citation distribution patterns and relatedness among sources.

## Results

The MEDLINE search retrieved 2083 records (see **Appendix 2** on OSF: https://osf.io/cpmgu/files/w4zha), of which 1966 records had DOIs; the subsequent DOI citation searches in Scopus yielded 1917 records (see **Appendix 3** on OSF: https://osf.io/cpmgu/files/e4xny). The present bibliometric analysis includes a total of 1917 publications (of which 921 were open-access), by 9749 unique authors, that were published between 1991 and 2025 (**Figure 1**). The five articles published in 2026 were available ahead of print. General characteristics of publications are presented in **Table 2**.

**Figure 1:**
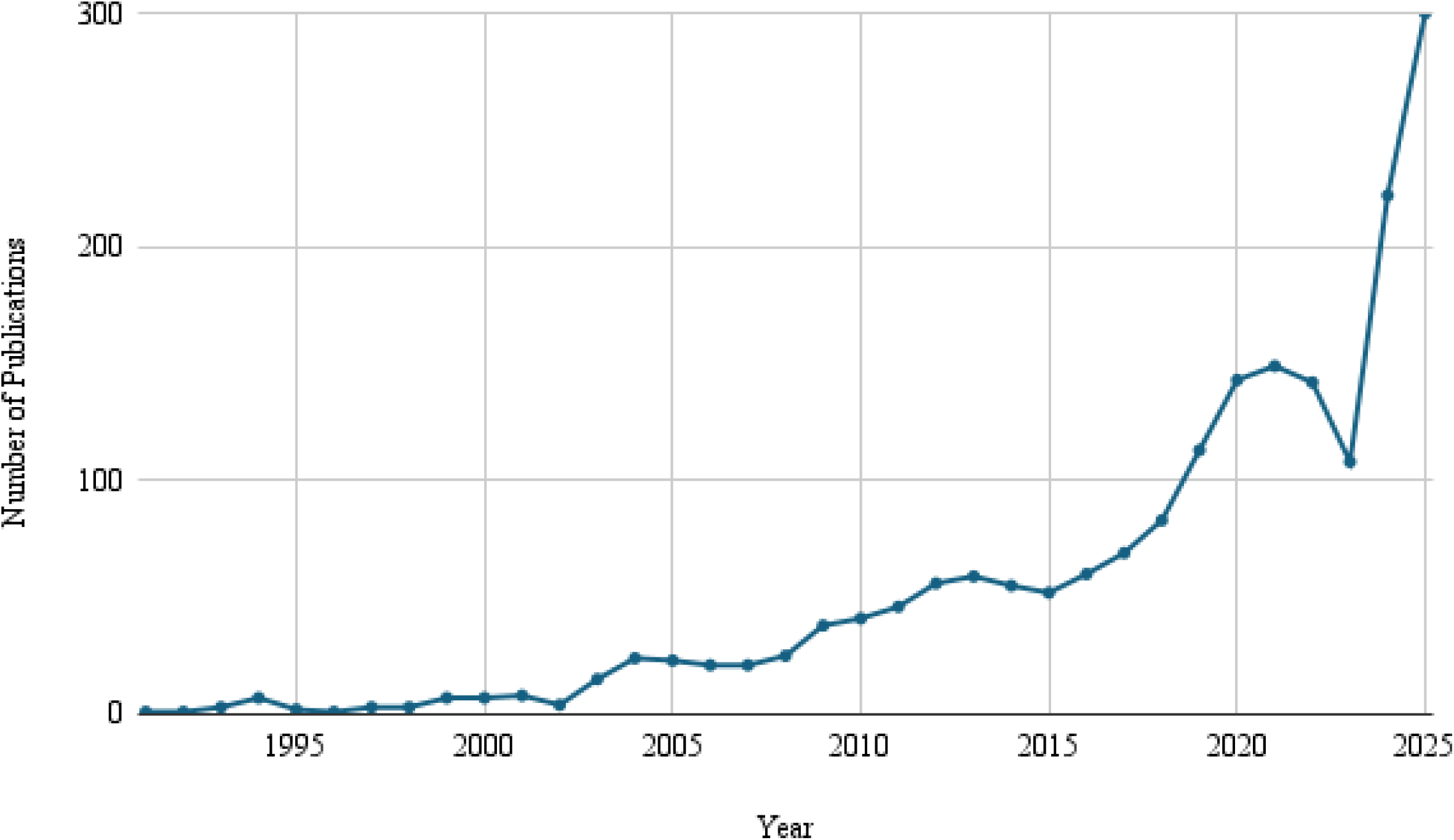
Annual Publication Volume from 1991 to 2025.

**Table 2:**
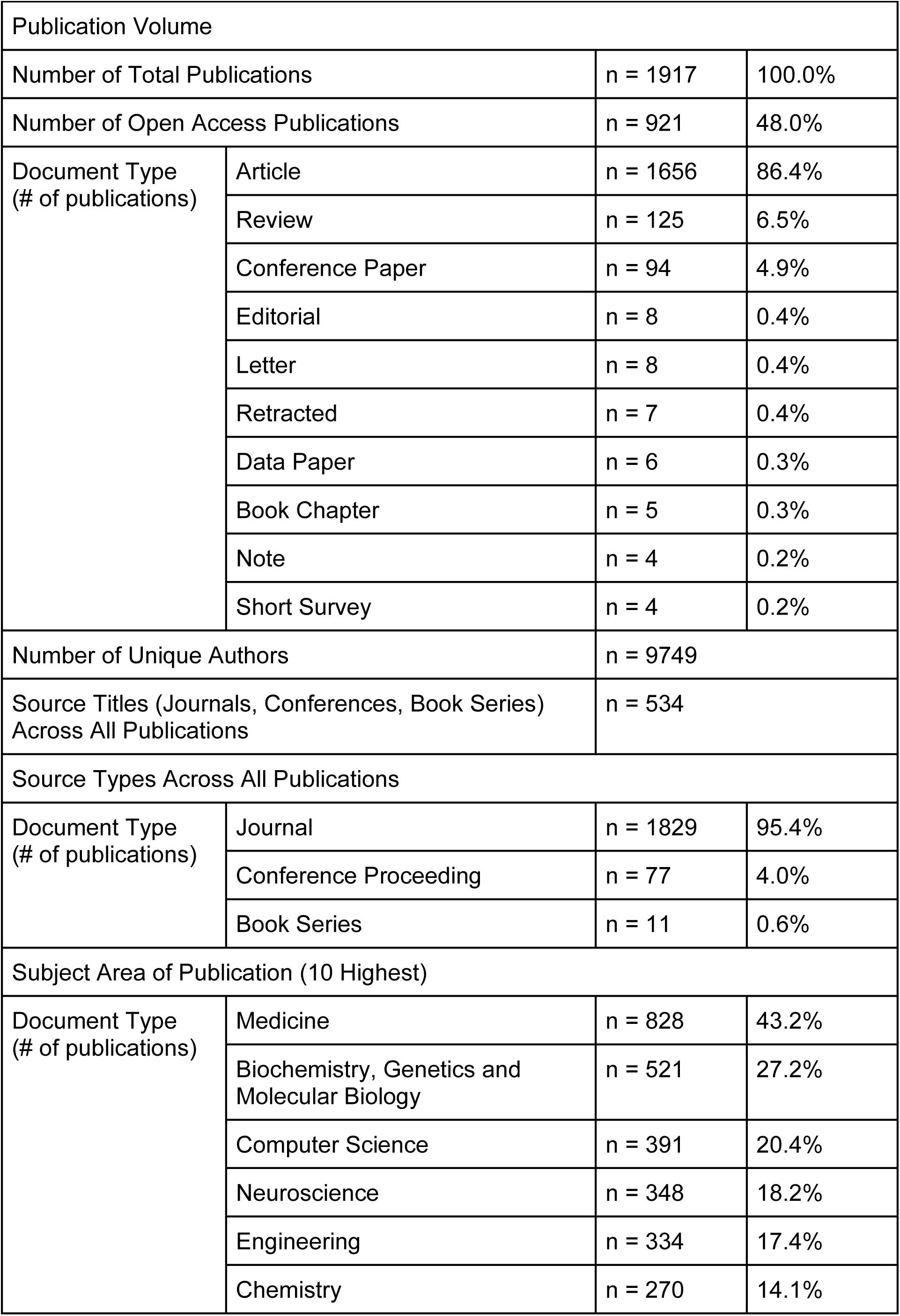
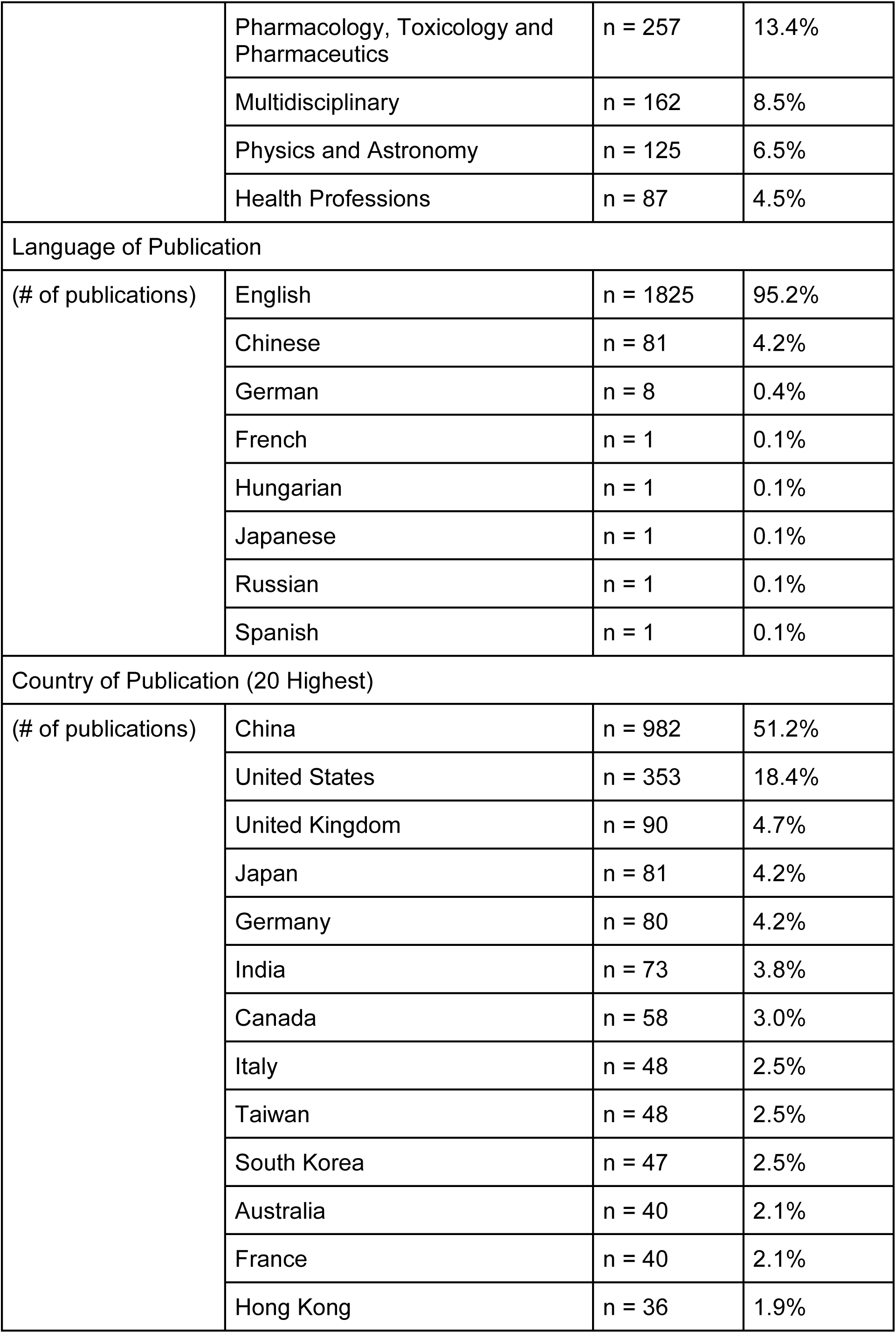
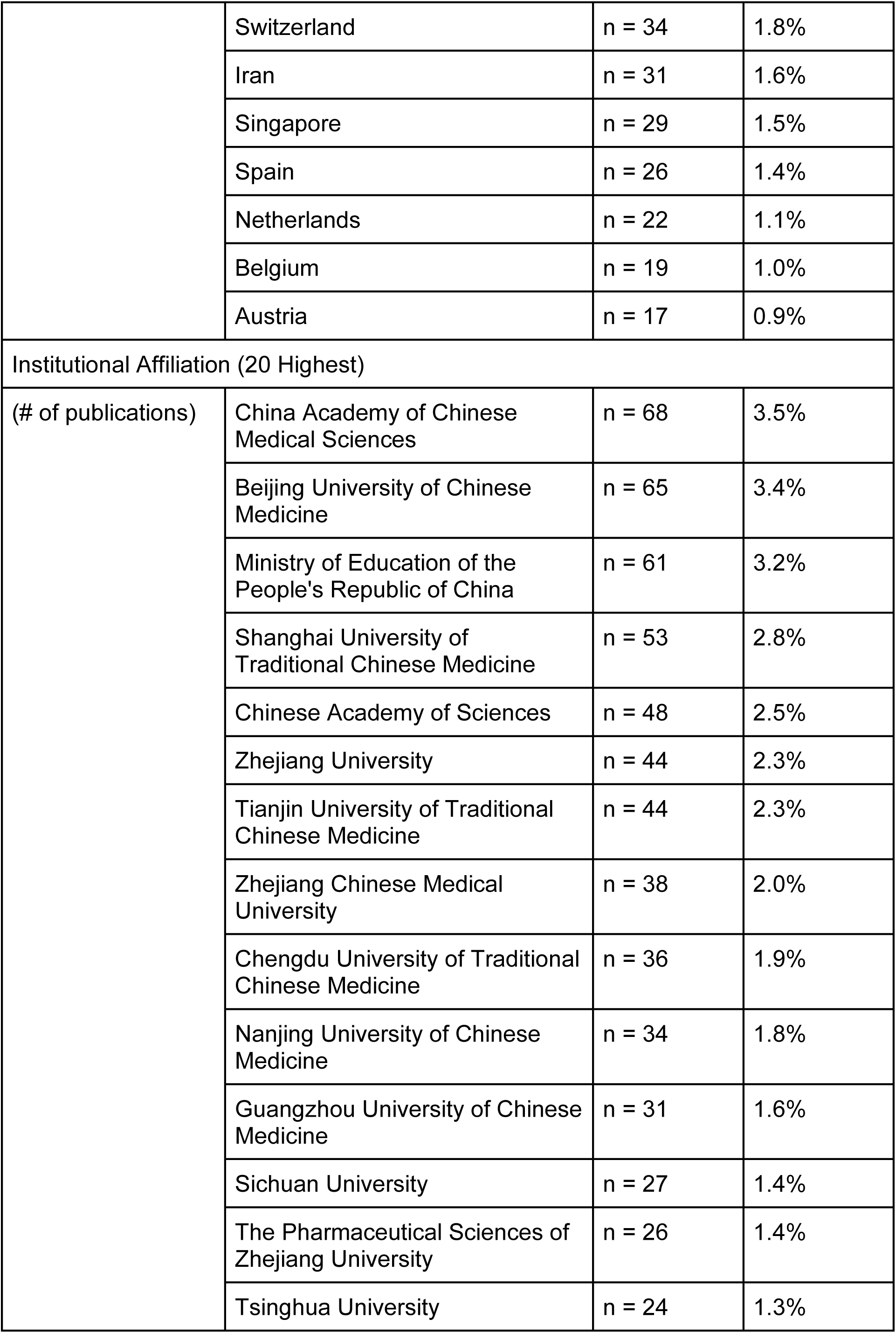
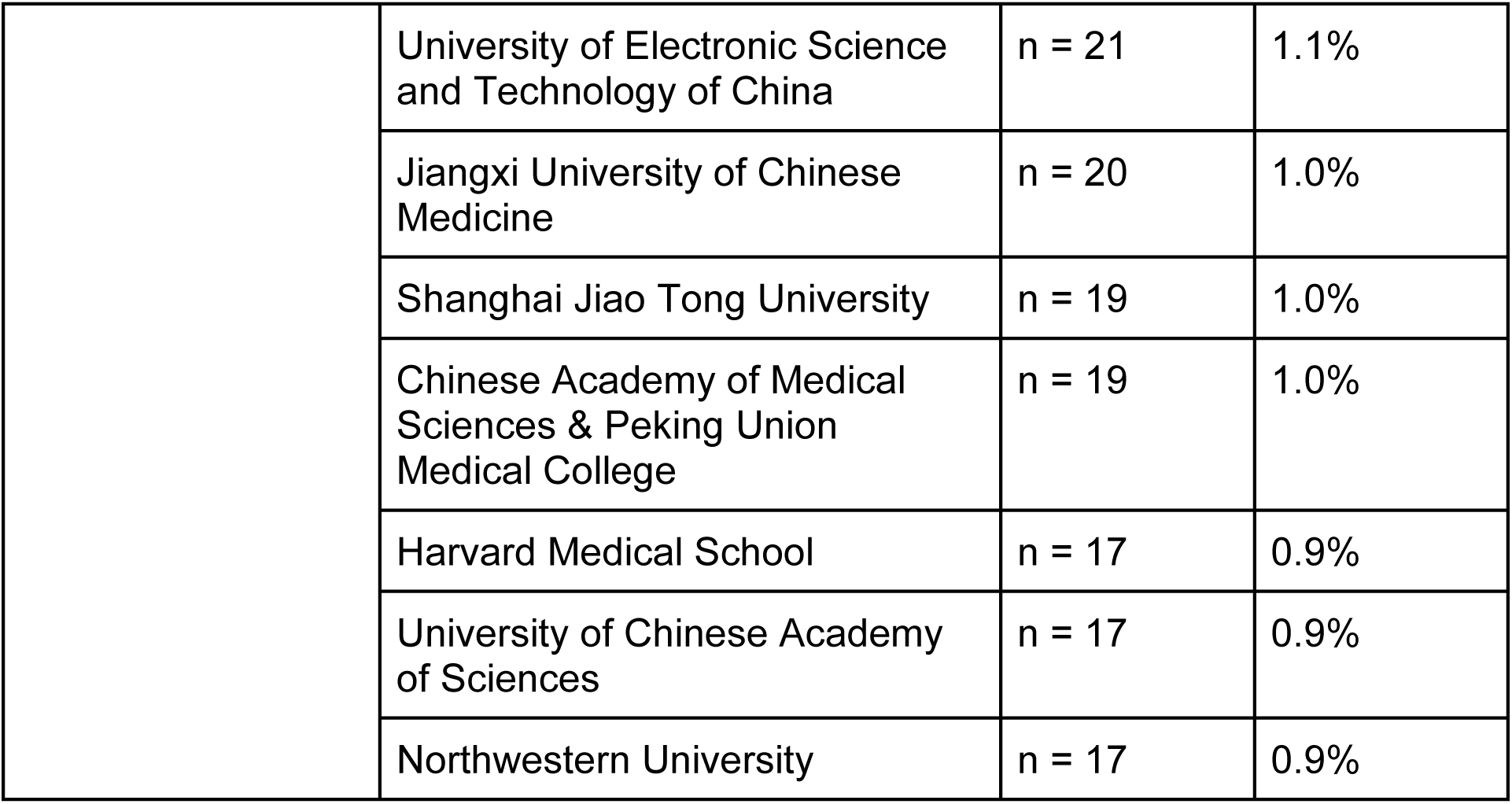
General Characteristics of Publications.

Publications were identified by Scopus as the following publication types: articles (n = 1656), reviews (n = 125), conference papers (n = 94), editorials (n = 8), letters (n = 8), retracted papers (n = 7), data papers (n = 6), book chapters (n = 5), notes (n = 4), and short surveys (n = 4). The vast majority of publication sources were journals (n = 1829), followed by conferences (n = 77) and book series (n = 11). The number of publications per source ranged from 1 to 86. Details regarding the top 10 sources with the highest number of publications are presented in **Table 3**, and the top 20 highest-cited publications are presented in **Table 4**.

**Table 3:**
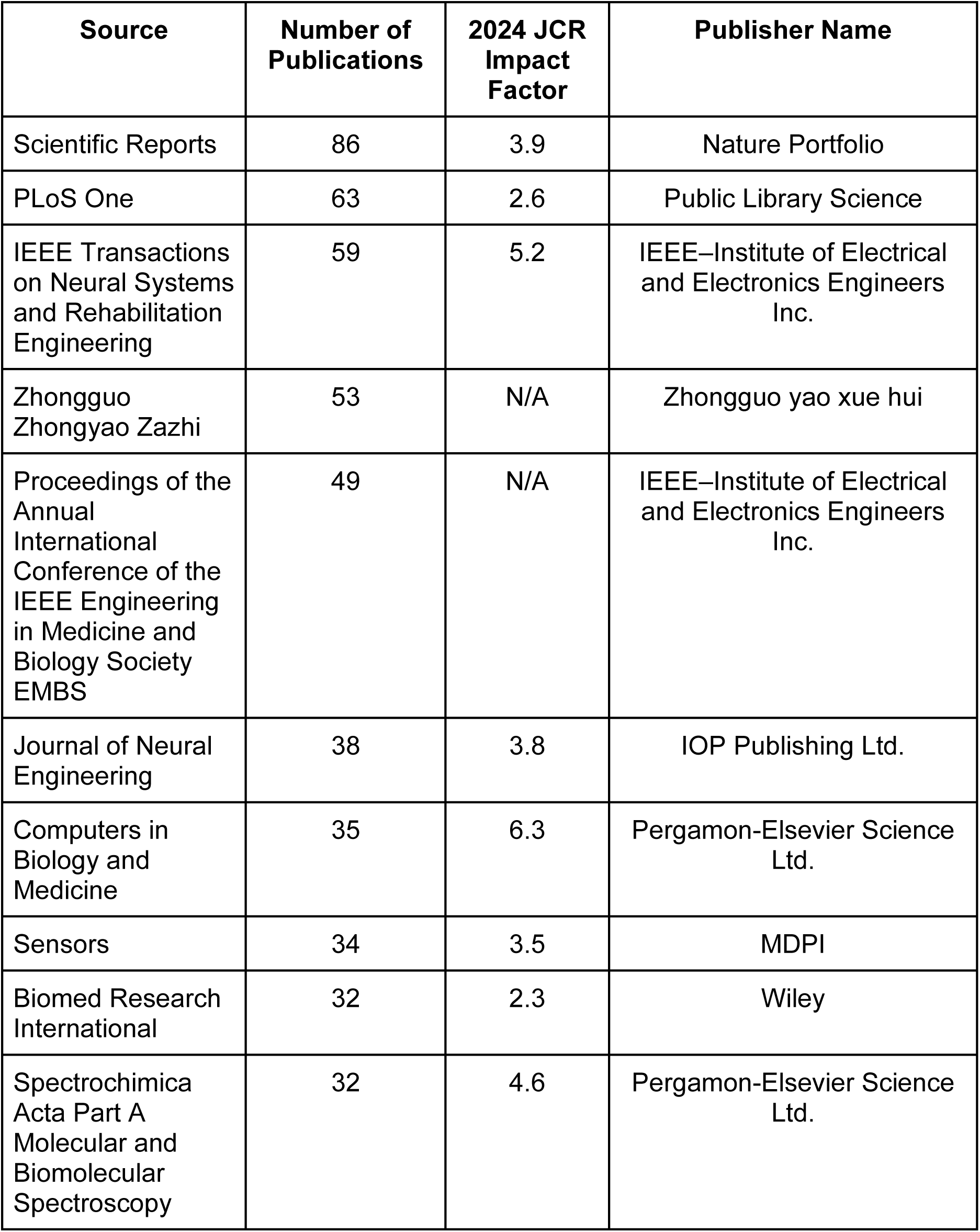
Characteristics of Top 10 Sources by Number of Publications.

| Source | Number of Publications | 2024 JCR Impact Factor | Publisher Name |
| --- | --- | --- | --- |
| Scientific Reports | 86 | 3.9 | Nature Portfolio |
| PLoS One | 63 | 2.6 | Public Library Science |
| IEEE Transactions on Neural Systems and Rehabilitation Engineering | 59 | 5.2 | IEEE–Institute of Electrical and Electronics Engineers Inc. |
| Zhongguo Zhongyao Zazhi | 53 | N/A | Zhongguo yao xue hui |
| Proceedings of the Annual International Conference of the IEEE Engineering in Medicine and Biology Society EMBS | 49 | N/A | IEEE–Institute of Electrical and Electronics Engineers Inc. |
| Journal of Neural Engineering | 38 | 3.8 | IOP Publishing Ltd. |
| Computers in Biology and Medicine | 35 | 6.3 | Pergamon-Elsevier Science Ltd. |
| Sensors | 34 | 3.5 | MDPI |
| Biomed Research International | 32 | 2.3 | Wiley |
| Spectrochimica Acta Part A Molecular and Biomolecular Spectroscopy | 32 | 4.6 | Pergamon-Elsevier Science Ltd. |

**Table 4:**
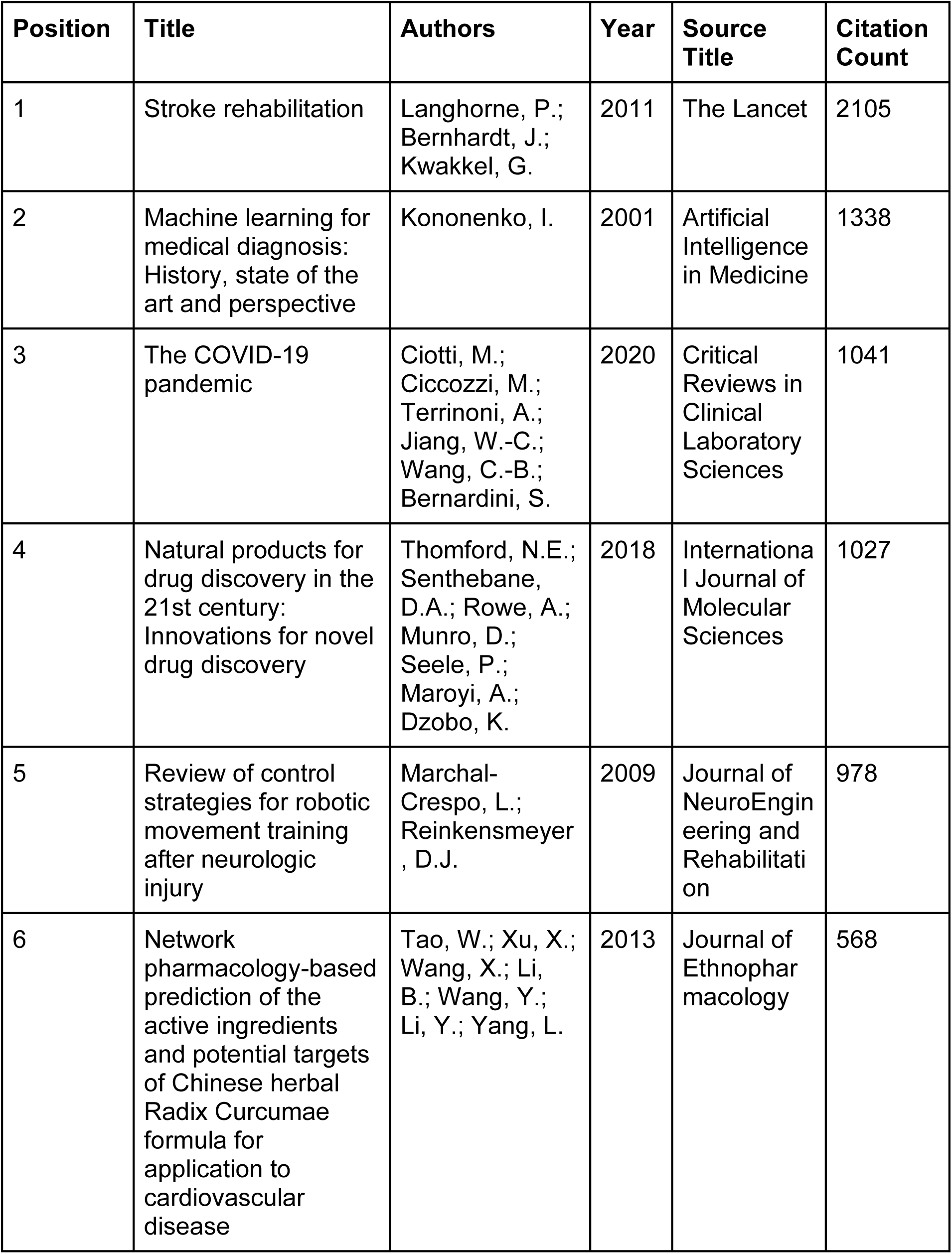
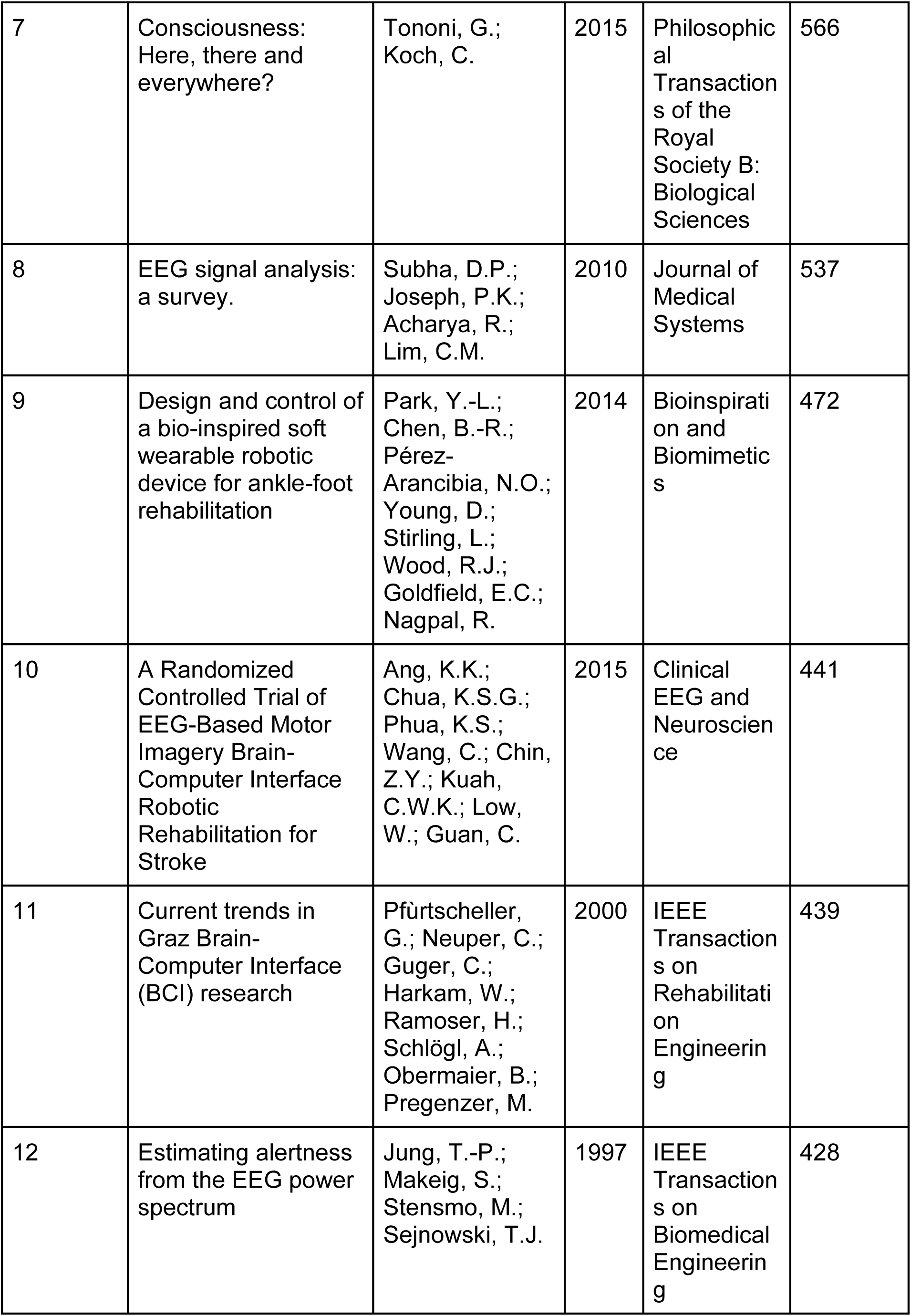
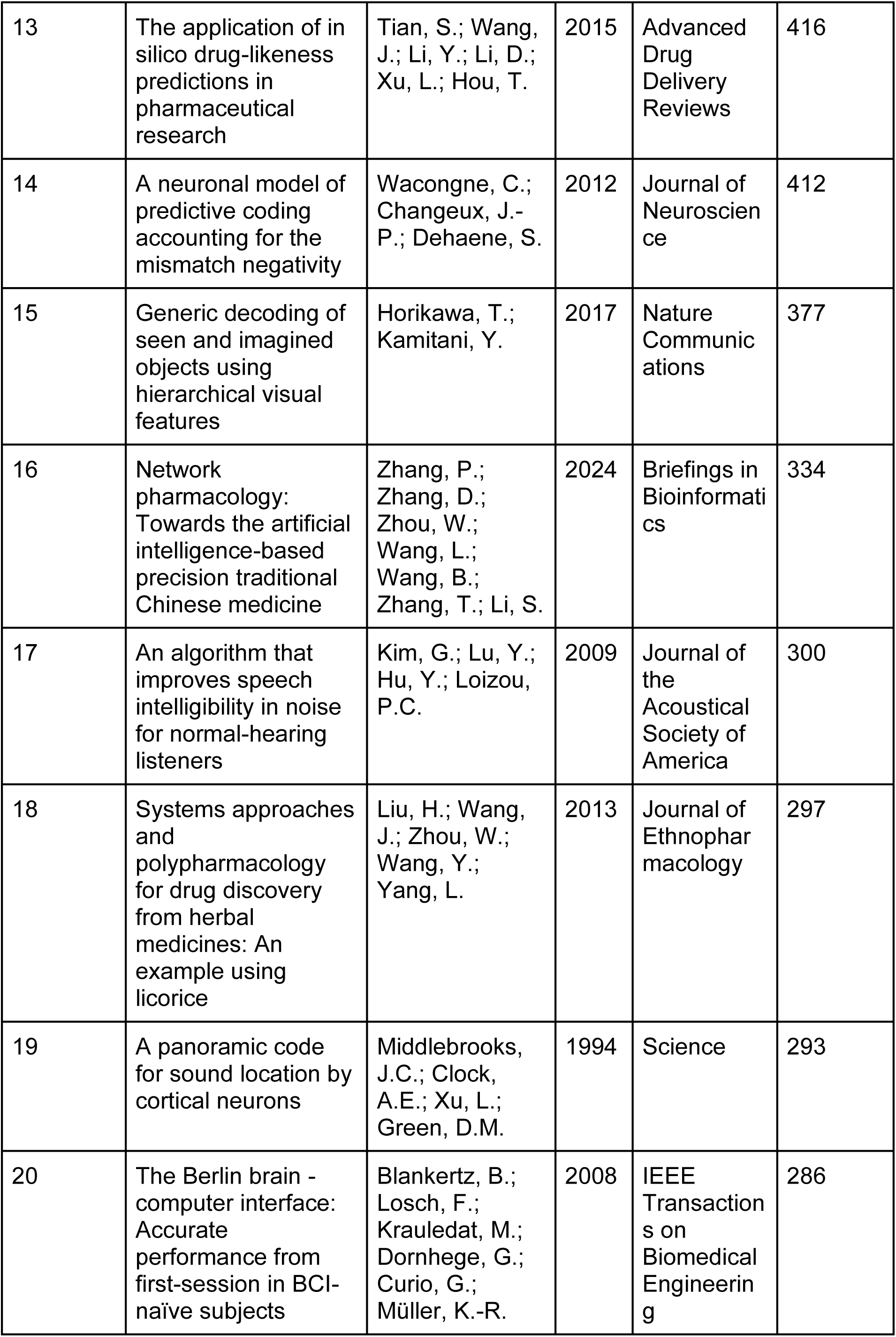
Characteristics of Top 20 Publications by Number of Citations.

The subject areas containing the largest number of publications were medicine (n = 828), followed by biochemistry, genetics and molecular biology (n = 521), computer science (n = 391), neuroscience (n = 348) and engineering (n = 334). Publication languages were primarily English (n = 1825), followed by Chinese (n = 81) and German (n = 8).

Authors were affiliated with 160 institutions across 86 countries. Publication volumes by region (**Figure 2**) were as follows: China (n = 982), the United States (n = 353), the United Kingdom (n = 90), Japan (n = 81), Germany (n = 80), India (n = 73), Canada (n = 58), Italy (n = 48), Taiwan (n = 48), South Korea (n = 47), Australia (n = 40), France (n = 40), Hong Kong (n = 36), Switzerland (n = 34), Iran (n = 31), Singapore (n = 29), Spain (n = 26), the Netherlands (n = 22), Belgium (n = 19), Austria (n = 17), Denmark (n = 17), Saudi Arabia (n = 15), Brazil (n = 14), Malaysia (n = 14), Pakistan (n = 13), Russia (n = 13), Thailand (n = 13), Finland (n = 11), Israel (n = 10), Poland (n = 10), and Sweden (n = 10). The remaining 55 countries had fewer than 10 publications (**Figure 3**).

**Figure 2:**
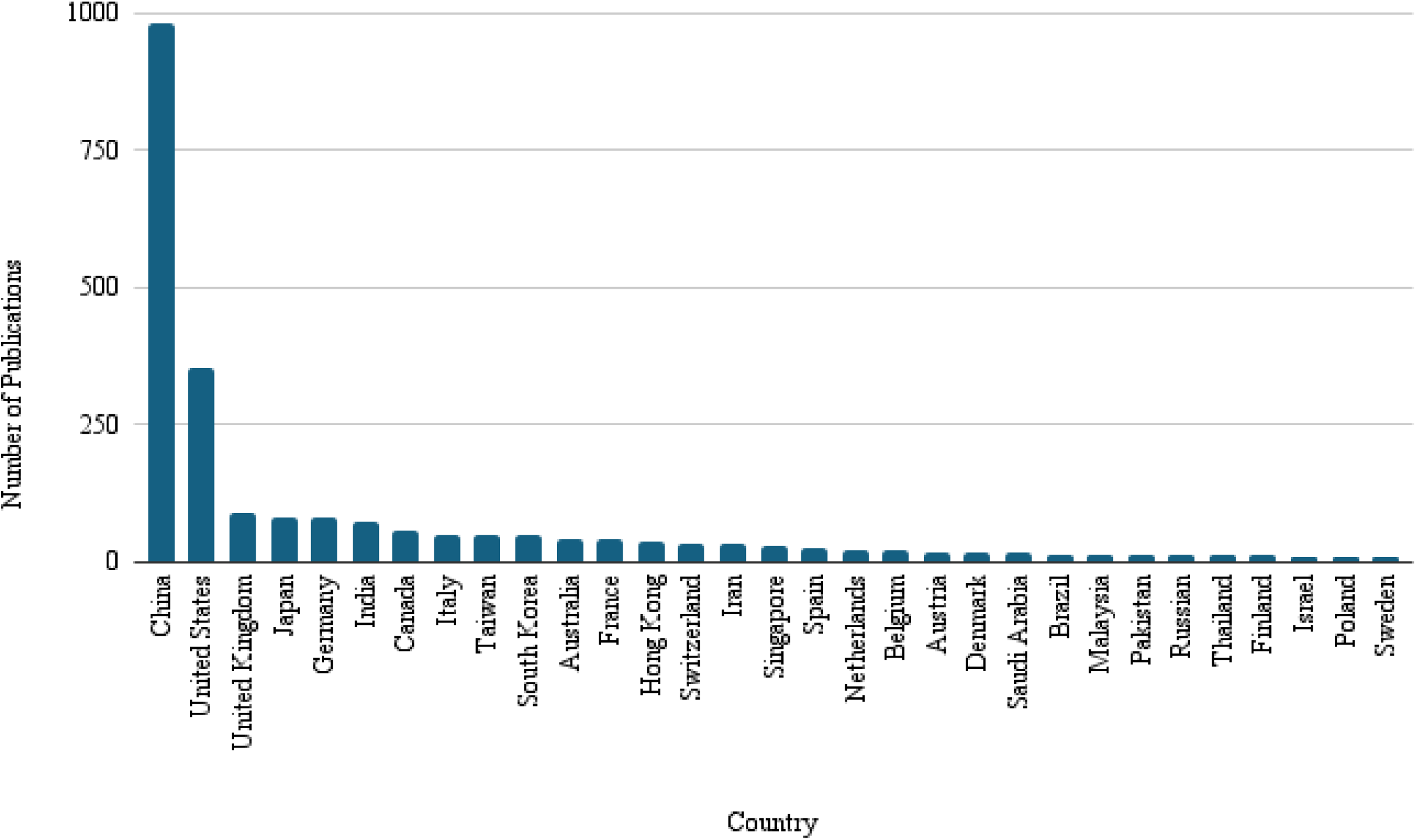
Total Number of Publications by Countries with 10 or More Publications.

**Figure 3:**
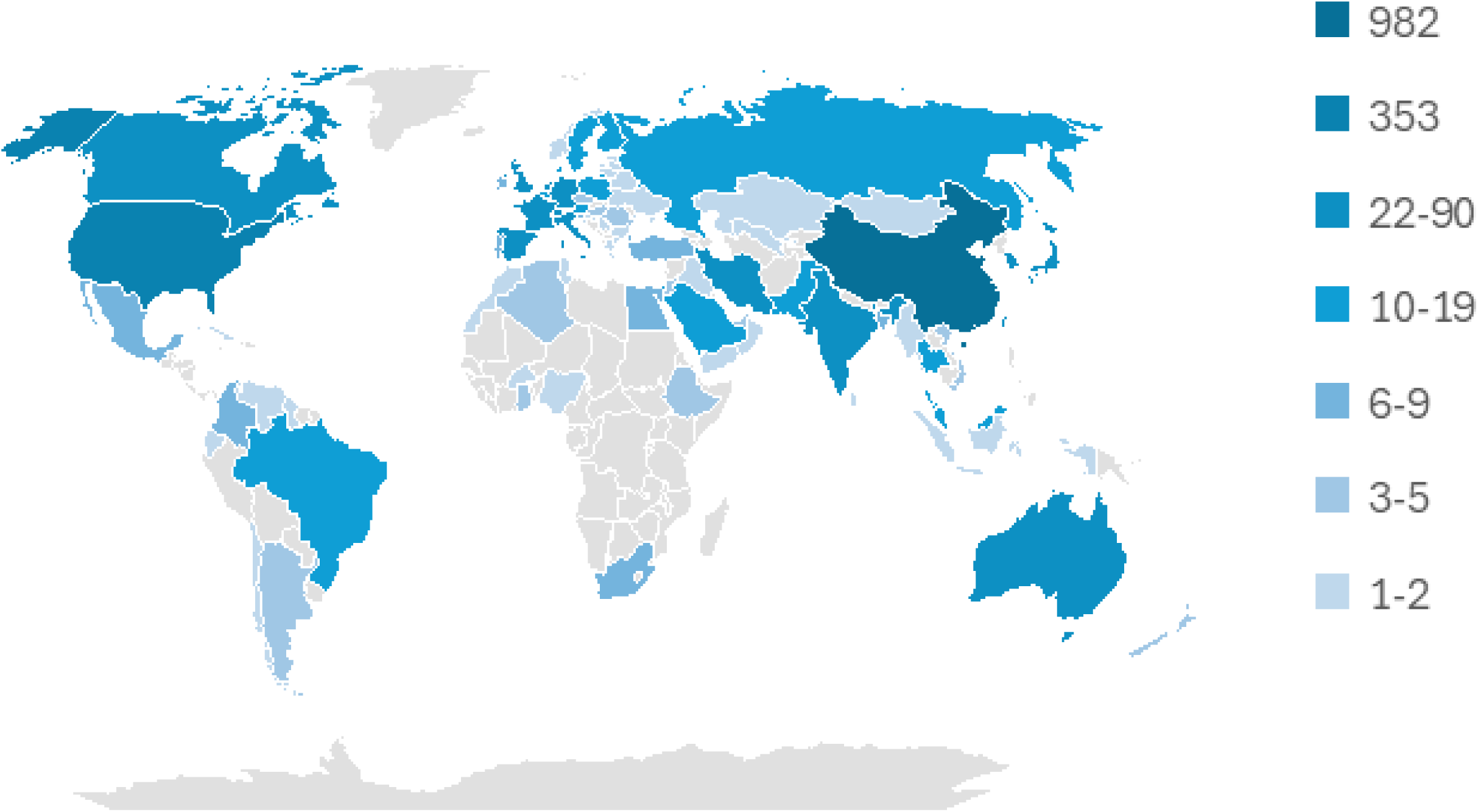
Distribution of Countries by Number of Publications.

The five most common affiliations were based in China and included the China Academy of Chinese Medical Sciences (n = 68), Beijing University of Chinese Medicine (n = 65), the Ministry of Education of the People’s Republic of China (n = 61), Shanghai University of Traditional Chinese Medicine (n = 53), and the Chinese Academy of Sciences (n = 48). Among the five highest funding sponsors, three institutions were based in China, including the National Natural Science Foundation of China (n = 380), the National Key Research and Development Program of China (n = 77), and the Fundamental Research Funds for the Central Universities (n = 37); and two institutions were based in the United States, including the United States National Institutes of Health (n = 65) and the National Science Foundation (n = 37).

**Figure 4** demonstrates a co-authorship analysis of the most productive countries with at least 10 publications each, where the links between countries is based on the number of co-authored publications.

**Figure 4:**
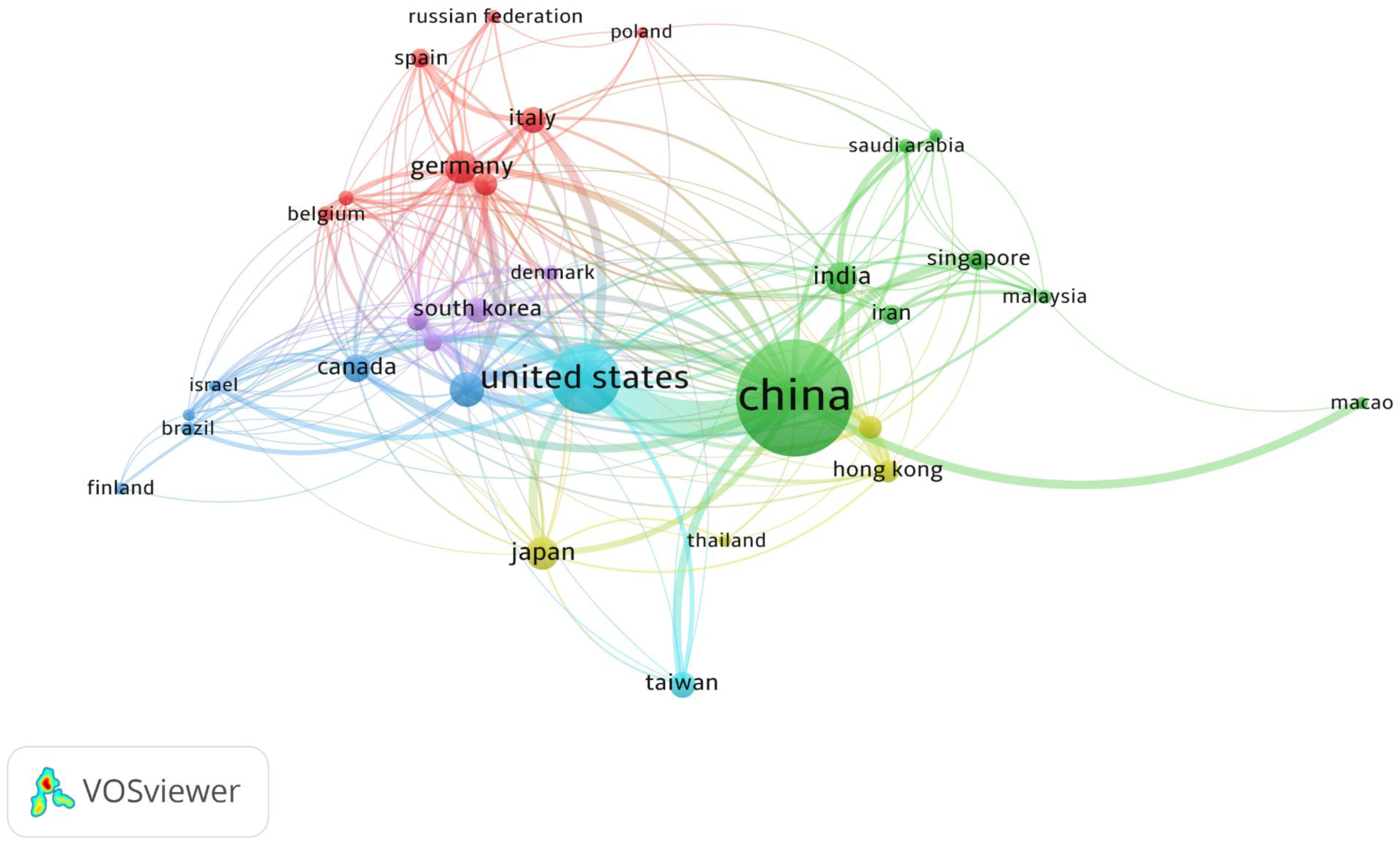
Co-Authorship Analysis of the Most Productive Countries with 10 or More Publications.

**Figure 5** demonstrates a co-occurrence analysis of the 100 most frequent author keywords used across all publications, where the links between keywords is based on the number of publications the keywords co-occur in together. The most frequent author keywords included “machine learning” (n = 182), “traditional Chinese medicine” (n = 127), “deep learning” (n = 100), “network pharmacology” (n = 97), “artificial intelligence” (n = 87), “EEG” (n = 43), “molecular docking” (n = 42), “stroke” (n = 37), “rehabilitation” (n = 32), “bioinformatics” (n = 28). Frequent TCIM-related author keywords included “traditional Chinese medicine” (n = 127), “acupuncture” (n = 18), “herbal medicine” (n = 16), “Chinese medicine” (n = 15), and “biofeedback” (n = 14); frequent AI-related author keywords included “deep learning” (n = 100), “artificial intelligence” (n = 87), “support vector machine” (n = 22), “classification” (n = 22), and “convolutional neural network” (n = 20). The 100 most frequent author keywords are provided in **Appendix 4** on OSF: https://osf.io/cpmgu/files/q8rn5.

**Figure 5:**
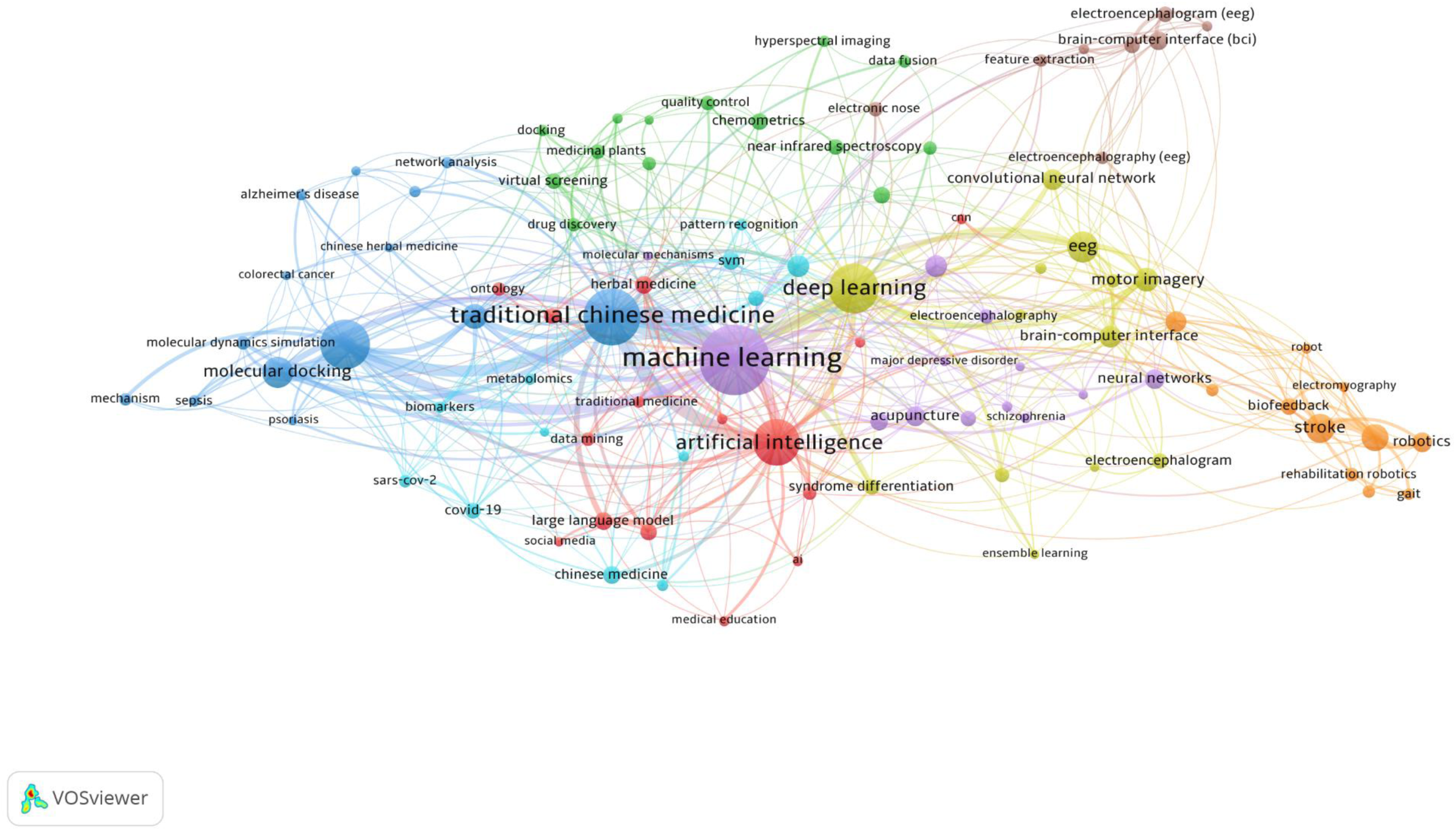
Co-Word Analysis of the 100 Most Frequent Author Keywords.

**Figure 6** demonstrates a co-citation analysis of the 100 most cited sources, where the links between sources is based on the frequency of sources being co-cited together. By total link strength, the most co-cited sources included the Journal of Neuroscience (n = 216), the Journal of Neurophysiology (n = 196), Nature (n = 183), Science (n = 139), and the Journal of the Acoustical Society of America (n = 119), the Proceedings of the National Academy of Sciences of the United States of America (n = 106), Neuroimage (n = 86), Hearing Research (n = 85), Clinical Neurophysiology (n = 83), and IEEE Transactions on Neural Systems and Rehabilitation Engineering (n = 77).

**Figure 6:**
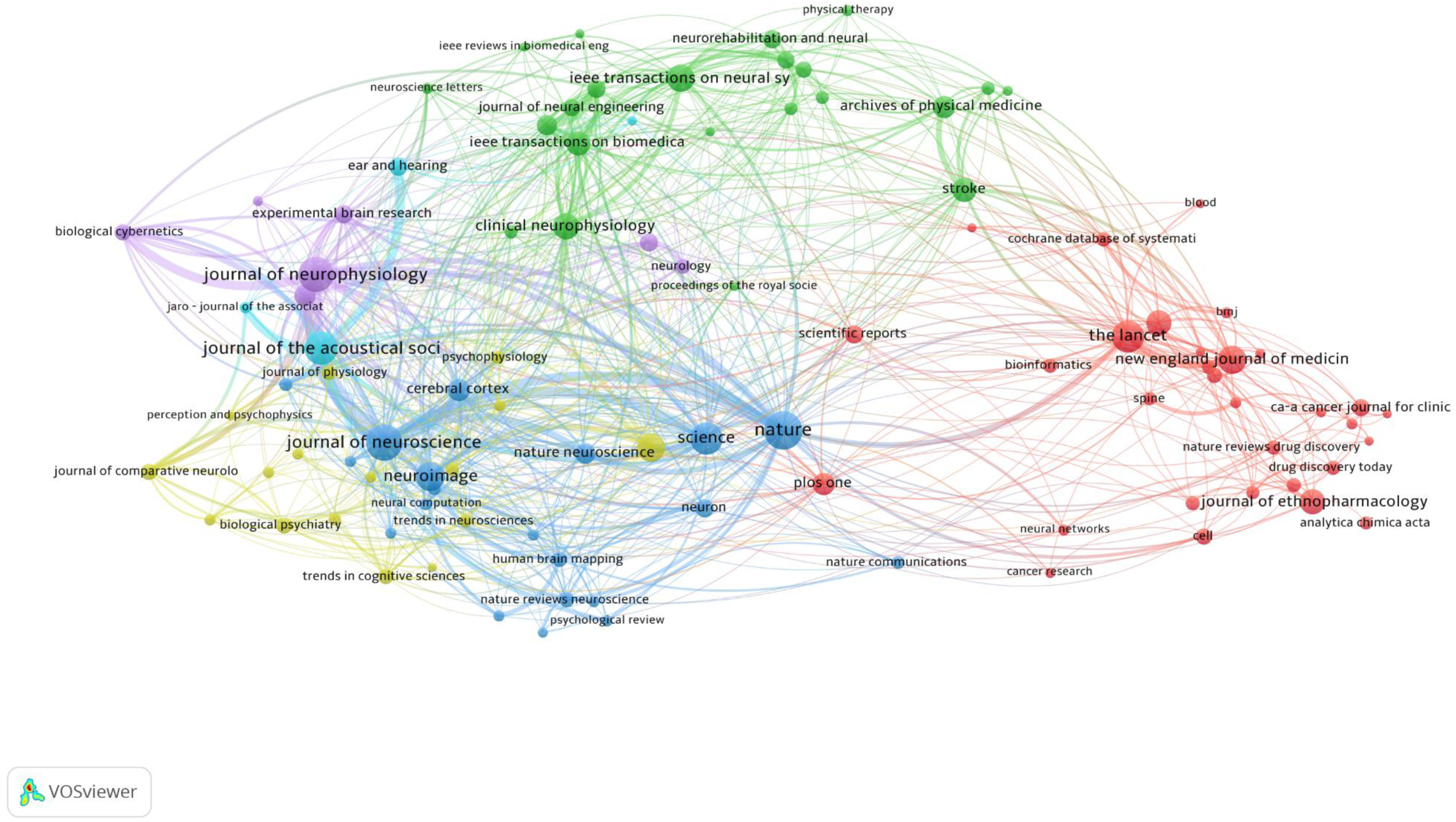
Co-Citation Analysis of the 100 Most Cited Sources.

## Discussion

This study systematically explores literature at the intersection of TCIM and AI fields by analyzing publication trends, key research contributors, citation impact, and keyword co-occurrences in the field. The database searches retrieved 1917 records for inclusion in the bibliometric analysis. A consistent upward trend since 2007 has been observed with respect to the annual number of publications, with the exception of a decrease in publications from 2021 to 2023, subsequently followed by a doubling in publication volume from 2023 to 2024. Over half of all articles have been published since 2020, mirroring the recent growth of research on AI in biomedicine generally.^34^ The top sources by number of publications included multidisciplinary open-access journals (e.g., Scientific Reports, PLoS One, Biomed Research International), Institute of Electrical and Electronics Engineers (IEEE) journals and conference proceedings, and other biotechnology journals (e.g., the Journal of Neural Engineering, Computers in Biology and Medicine). Notably, only one of the top 10 sources was a TCIM-specific journal (i.e., Zhongguo Zhongyao Zazhi).

In the co-authorship analysis (Figure 4), authors from China tend to have more international collaborations with many other countries, as did authors from the United States and the United Kingdom. The greatest degree of inter-country collaborations occurred between China and the United States. By contrast, German authors tend to collaborate more with European countries, and Japanese authors tend to collaborate more with East Asian countries.

Given the predominance of Chinese authors, institutions, and affiliations, it is unsurprising that frequent TCIM-related author keywords included “traditional Chinese medicine”, “acupuncture”, and “Chinese medicine”. Interestingly, however, specific TCIM therapies were associated with different keywords in the co-word analysis (Figure 5). For instance, in the dark blue cluster, “traditional Chinese medicine” was associated with drug discovery-related terms of “network pharmacology”, “systems pharmacology”, and “molecular docking”. Similarly, the red cluster paired AI- and language-related keywords of “large language model” and “natural language processing” with TCIM keywords of “traditional medicine” and “herbal medicine”. The purple cluster comprised AI terms of “machine learning” and “neural networks” with TCIM therapies like "acupuncture".

It can also be observed in the co-word analysis that some of the more studied disease states included cardiovascular conditions (i.e., coronary heart disease, stroke), infections and dysregulated immunities (i.e., COVID-19, sepsis, psoriasis), and psychiatric and neurological conditions (i.e., schizophrenia, spinal cord disease, Alzheimer’s disease), highlighting the broad applicability of AI and TCIM research. By comparison, these conditions differed from the findings of previous bibliometric analyses of TCIM, which found the most frequent disease-related keywords to include breast cancer, lung cancer, diabetes, anxiety, and low back pain.^33^

In the co-citation analysis (Figure 6), the dark blue, purple, and yellow clusters were characterized by closer association of neurosciences and psychology journals being more frequently co-cited together. The light blue cluster included a few audiological journals, and the green cluster mainly comprised IEEE sources, physical therapy journals, and rehabilitation journals. The red cluster was less well-defined, comprising high-impact general biomedical journals (e.g., Lancet, JAMA, BMJ), multidisciplinary open-access journals (e.g., Scientific Reports, PLoS One), TCIM journals (e.g., Journal of Natural Products, Evidence-Based Complementary and Alternative Medicine), and various other subject-specific biomedical journals (e.g., Circulation, Blood, Spine, Cancer Research, Bioinformatics, Hepatology).

### Comparative Literature

While there are, to the best of the authors’ knowledge, no previous bibliometric analyses examining research productivity at the convergence of the AI field and the TCIM field as a whole, other bibliometric studies have analyzed the intersection of AI and specific TCIM therapies (e.g., TCM, acupuncture, traditional medicine).^40–42^ In comparison to the existing literature on TCIM therapies and AI, this bibliometric analysis shared some similar broad trends: the volume of publications generally demonstrated upwards growth patterns, the most productive countries included China and the United States, and the most common institutional affiliations frequently included Chinese and American universities and organizations.^40–42^

Cao et al. (2024)^40^ examined the research landscape of AI in TCM, identifying 1183 research articles and reviews by 6626 authors published from 2000 to 2024. This bibliometric analysis described an exponential growth trend in annual publications, with a surge in growth since 2013. The most productive countries were identified to be China (n = 643), the United States (n = 160), and India (n = 60). Accordingly, the most common institutional affiliations included the China Academy of Chinese Medical Sciences (n = 39), Beijing University of Chinese Medicine (n = 36), and Shanghai University of Traditional Chinese Medicine (n = 35).^39^ The most productive journals were identified as Molecules (n = 26), Scientific Reports (n = 25), Evidence-Based Complementary and Alternative Medicine (n = 25), and IEEE Access (n = 25). The most frequently co-occurring keywords included machine learning, traditional Chinese medicine, and deep learning, while the most frequently mentioned disease states included hepatocellular carcinoma, chemical and drug-induced liver damage, Papillon-Lefèvre disease, Parkinson’s disease, and anorexia.^40^

Zhou et al. (2023)^41^ analyzed research on AI in acupuncture, finding 417 publications from 1994 to 2022, also identifying an exponential growth trend in annual publications. The countries with the highest number of publications were the United States (n = 154), China (n = 85), and Japan (n = 38); however, by average citations, the highest-ranking nations were France (n = 56.69), Canada (n = 47.32), and the United Kingdom (45.59). The most frequent institutional affiliations were Harvard University (n = 15), University of California System (n = 11), Harvard Medical School (n = 10), and Seoul National University (n = 10). The most prolific journals included the Journal of Alternative and Complementary Medicine (n = 10), Diagnostic Cytopathology (n = 7), Diagnostics (n = 6), Evidence-Based Complementary and Alternative Medicine (n = 6), and IEEE Robotics and Automation Letters (n = 6).^41^

Liu et al. (2024)^42^ investigated applications of ML in traditional medicine, analyzing 282 English-language publications by 1601 authors from 2012 to 2022. Although generally demonstrating a growth trend, the volume of publications peaked in 2020 with 78 papers. The most prolific countries were China (n = 152), the United States (n = 92), and England (n = 20). The most frequent affiliations were Shanghai University of Traditional Chinese Medicine (n = 19), University of California, San Francisco (n = 19), and University of Toronto (n = 16). The most productive journals were IEEE Access (n = 15); Evidence-Based Complementary and Alternative Medicine (n = 7), and JMIR Medical Informatics (n = 6).^42^

### Strengths and Limitations

A key strength of this study lies in its comprehensive and systematic approach to identifying and analyzing the intersection of TCIM and AI research. The use of MEDLINE as the primary data source ensured broad coverage of peer-reviewed literature in biomedical and life sciences. The application of advanced bibliometric techniques, including co-authorship, co-citation, and co-word analysis, using VOSviewer provided a multidimensional view of the research landscape. Furthermore, this study’s registration to OSF ensured methodological transparency and reproducibility. However, certain limitations should be acknowledged. Database coverage bias may have been present, as MEDLINE does not index all TCIM or AI-related journals or articles, potentially omitting relevant records. Language biases may also have affected results, as non-English publications that are not translated could be underrepresented. Further, given the procedure of DOI citation searches in Scopus, differences in database coverage between MEDLINE and Scopus resulted in omission of retrieved records that are indexed by MEDLINE but not Scopus. Similarly, MEDLINE records that lack DOIs were also excluded from the dataset. A further limitation was the lack of manual screening of results, where out-of-scope articles could potentially have been included in this bibliometric analysis. Finally, bibliometric analyses do not assess the quality or validity of included research; thus, findings should be interpreted as indicators of publication trends rather than definitive measures of scientific impact.

## Conclusion

The present study examined the characteristics of literature at the intersection of AI and TCIM fields, identifying 1917 publications from 1991 to 2026, with a steep rise in the volume of publications since 2023. The most productive countries included China, the United States, and the United Kingdom; correspondingly, a great proportion of the most productive institutional affiliations and funding sponsors were from these countries. Acupuncture, TCM, and herbal medicine are among the more researched therapies leveraging the use of AI technologies in research areas such as drug discovery. Given the increasing use of TCIM among patients and medical professionals, as well as growing integration of AI into healthcare systems, other relatively underexplored TCIM therapies (e.g., Ayurvedic medicine, osteopathic medicine, mind-body therapies) can be areas of future research.

## Data Availability

All relevant data and materials are provided in this manuscript or are posted on the Open Science Framework.

https://doi.org/10.17605/OSF.IO/CPMGU

## List of Abbreviations

AI: artificial intelligence
DL: deep learning
DOI: digital object identifier
JCR: Journal Citation Reports
IEEE: Institute of Electrical and Electronics Engineers
MeSH: Medical Subject Headings
ML: machine learning
NLP: natural language processing
OSF: Open Science Framework
TCIM: traditional, complementary, and integrative medicine
TCM: traditional Chinese medicine
WHO: World Health Organization

## Declarations

### Ethics Approval and Consent to Participate

This study involved a bibliometric analysis of the literature only; the study did not require ethics approval or consent to participate.

### Consent for Publication

All authors consent to this manuscript’s publication.

### Availability of Data and Materials

All relevant data and materials are provided in this manuscript or posted on the Open Science Framework at https://doi.org/10.17605/OSF.IO/CPMGU.

### Authors’ Contributions

HL: co-drafted the manuscript, collected and analysed data, and gave final approval of the version to be published.

M-IB: designed and conceptualized the study, collected and analysed data, co-drafted the manuscript, and gave final approval of the version to be published. JYN: designed and conceptualized the study, collected and analysed data, co-drafted the manuscript, and gave final approval of the version to be published.

